# “Injury-Prone”: Peripheral nerve injuries associated with prone positioning for COVID-19-related acute respiratory distress syndrome

**DOI:** 10.1101/2020.07.01.20144436

**Authors:** George R. Malik, Alexis R. Wolfe, Rachna Soriano, Leslie Rydberg, Lisa F. Wolfe, Swati Deshmukh, Jason H. Ko, Ryan P. Nussbaum, Prakash Jayabalan, James M. Walter, Colin K. Franz

**Affiliations:** McGaw Medical Center, Northwestern University Feinberg School of Medicine, Chicago, IL, USA; Shirley Ryan Ability Lab (Formerly the Rehabilitation Institute of Chicago), Chicago, IL, USA; Department of Physical Medicine and Rehabilitation, Northwestern University Feinberg School of Medicine, Chicago, IL, USA; The Division of Pulmonary and Critical Care, Department of Medicine, Northwestern University Feinberg School of Medicine, Chicago, IL, USA; The Ken and Ruth Davee Department of Neurology, Northwestern University Feinberg School of Medicine, Chicago, IL, USA; Department of Radiology, Northwestern University Feinberg School of Medicine, Chicago, IL, USA; Division of Plastic and Reconstructive Surgery, Northwestern University Feinberg School of Medicine, Chicago, IL, USA

## Abstract

Patients with Coronavirus disease 2019 (COVID-19) who require invasive mechanical ventilation frequently meet the acute respiratory distress syndrome (ARDS) diagnostic criteria. Hospitals based in the United States have been incorporating prone positioning (PP) into the COVID-19-related ARDS treatment plan at a higher rate than normal. Here, we describe 11 patients admitted to a single inpatient rehabilitation hospital who were subsequently diagnosed with acquired focal/multifocal peripheral nerve injury (PNI) in association with the use of PP for COVID-19-related ARDS. The reason for the high rate of PNI associated with PP in COVID-19 ARDS is likely multifactorial, but may include an underlying state of hyperinflammation and hypercoagulability already linked to other the neurological sequelae of COVID-19. Physicians must be aware of this elevated susceptibility to PNI in severe COVID-19 and refined standard PP protocols in order to reduce the risk.

## Introduction

Patients with Coronavirus disease 2019 (COVID-19) who require invasive mechanical ventilation frequently meet the acute respiratory distress syndrome (ARDS) diagnostic criteria. Both guidelines and expert opinion recommend 12 to 16 hours per day of prone positioning (PP) for patients with moderate-to-severe ARDS from COVID-19 [1]. Many of the most severely affected survivors from COVID-19 are now being discharged to inpatient rehabilitation (IPR) hospitals [2]. This provides an important opportunity to assess the long-term sequelae of PP in this unique patient population. Here, we describe 11 patients who were diagnosed with acquired focal/multifocal peripheral nerve injury (PNI) in association with the use of PP for COVID-19-related ARDS.

## Methods

Study approval was granted by the Northwestern University Institutional Review Board. Patients were identified during their admission to a single IPR hospital (Shirley Ryan AbilityLab, Chicago, IL). Clinical suspicion of an acquired PNI was based on history and physical examination performed on admission. Use of PP while patients were mechanically ventilated was confirmed either through review of the medical record or, when records were not available, interview with family members. Whenever possible the diagnosis of PNI was made by electromyography and nerve conduction studies (EMG/NCS). In one case, the localization of PNI was aided by magnetic resonance neurography (**Figure 1a**). In another case, a patient was discharged without EMG/NCS test completed due to the temporary closure of the electrodiagnostic laboratory early in COVID-19 pandemic, so the diagnosis of PNI was made by physical exam. Mean data values are expressed as plus or minus (±) standard deviation.

**Figure 1.**
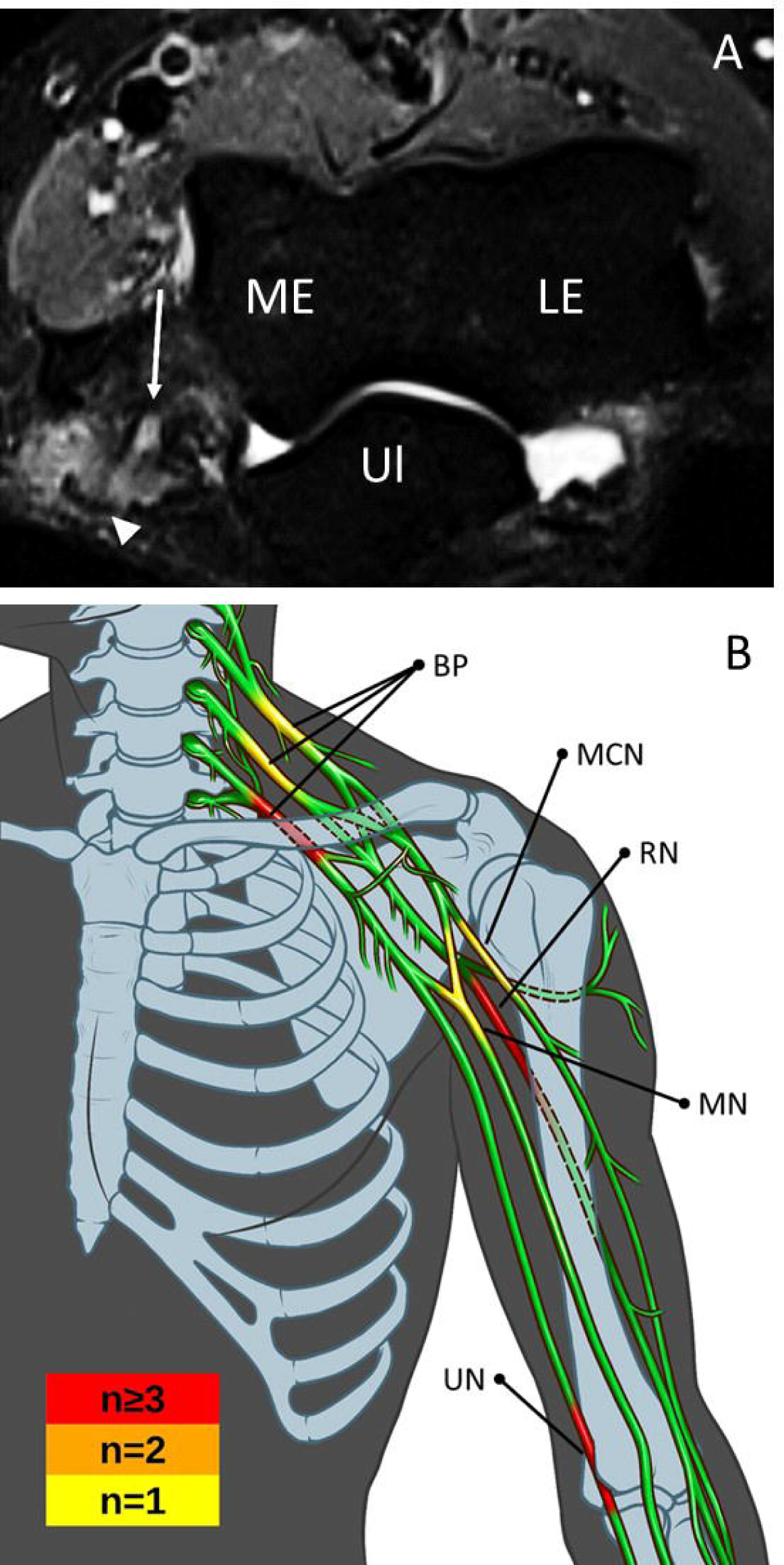
Locations of upper limb peripheral nerve injuries associated with prone positioning of patients with COVID-19-related ARDS. A, Axial STIR image from MR neurography of the left humerus (patient 5) demonstrates focal signal hyperintensity of the ulnar nerve (arrow) with focal adjacent soft tissue edema (arrowhead). B, Graphical summary of all upper limb PNI sites in this report. Heat map represents the frequency of PNI at defined anatomical sites. ME, medial epicondyle; LE, lateral epicondyle; Ul, ulna; BP, brachial plexus; MCN, musculocutaneous nerve; RN, radial nerve; MN, median nerve; UN, ulnar nerve.

## Results

Between 4/24/2020 and 6/30/2020, 85 patients were admitted to our IPR facility following hospitalization for COVID-19-related ARDS. Of these, 11 patients (12.9%) were then diagnosed with PNI. Demographic and clinical characteristics for these 11 patients are provided (**Table 1**). Patients in our cohort received their care at six different acute care hospitals. All patients had received PP while mechanically ventilated for COVID-19-related ARDS. The number of PP sessions was 4.5 +/- 5.6 per patient and the number of hours spent in the prone position was 81.2 +/- 89.6 hours. The total duration of invasive mechanical ventilation was 34.9 +/- 15.0 days. In total, there were 18 separate PNIs across these 11 patients (1.6 ± 1.0 PNIs/patient). The vast majority of these newly diagnosed PNIs occurred in the upper limb (83.3%; **Figure 1**). The most frequent sites of injury included the ulnar nerve (5 cases, 27.7%), radial nerve (3 cases, 16.6%), and the lower trunk of the brachial plexus (3 cases, 16.6%). All patients had axontomesis type PNI based on Seddon classification system, which ranged from moderate to severe in terms of reduction in motor unit recruitment parameter on EMG/NCS.

**Table 1.** Summary of patient demographics and key clinical characteristics. HLD, hyperlipidemia; DM2, type II diabetes mellitus; HTN, hypertension; Pre-DM2, pre-type II diabetes mellitus; COPD, chronic obstructive pulmonary disease; DM1, type I diabetes mellitus.

| Patient | Age | Sex | Ethnicity | BMI | Comorbidities | EMG | Location | Grade | Days of mechanical ventilation | Proning sessions (#) | Hours spent prone (total) | Days of neuromuscular blockade |
| --- | --- | --- | --- | --- | --- | --- | --- | --- | --- | --- | --- | --- |
| 1 | 72 | M | Caucasian | 30 | HLD | Y | Right radial n., Right ulnar n. | Severe | 62 | 2 | 35 | 3 |
| 2 | 66 | F | Caucasian | 37 | HLD, DM2 | Y | Right radial, Right middle trunk plexus, Right lower trunk plexus | Severe | 43 | 2 | 32 | NA |
| 3 | 68 | M | African American | 43 | HTN, HLD | Y | Right lower trunk plexus | Moderate | 38 | 4 | 63 | 8 |
| 4 | 74 | F | Caucasian | 50 | HTN, HLD, DM2 | X | Left radial n. | N/A (no EMG) | 12 | 1 | 16 | 1 |
| 5 | 45 | M | Hispanic | 27 | HLD, Pre-DM2 | Y | Right ulnar n., Left ulnar n., Right musculocutaneous n., Left median n. | Severe | 25 | Unknown | Unknown | 5 |
| 6 | 60 | M | Hispanic | 26 | HTN, DM2 | Y | Right ulnar n. | Severe | 46 | 1 | Unknown | Unknown |
| 7 | 57 | M | Caucasian | 30 | HTN, HLD, DM2 | Y | Left sciatic n. | Severe | 47 | 16 | 192 | Unknown |
| 8 | 55 | M | African American | 25 | HTN, Asthma | Y | Right upper trunk plexus | Moderate | Unknown | 1 | 16 | Unknown |
| 9 | 63 | M | Hispanic | 31 | HTN | Y | Right ulnar n., Right sciatic n. | Severe | 31 | 14 | 252 | 1 |
| 10 | 80 | F | African American | 34 | HTN, COPD, DM2 | Y | Right lower trunk plexus | Moderate | 21 | 3 | 44 | 1 |
| 11 | 23 | F | Hispanic | 28 | DM1 | Y | Right deep fibular n. | Severe | 24 | 1 | Unknown | 17 |
| MEAN | 60.3 |  |  | 32.8 |  |  |  |  | 34.9 | 4.5 | 81.2 | 5.1 |
| SD | 15.7 |  |  | 7.8 |  |  |  |  | 15.0 | 5.6 | 89.6 | 5.8 |

## Discussion

This report represents the single largest description of new PNIs associated with PP for management of ARDS, and to our knowledge, the first description specific to COVID-19. PNIs are a well-known, albeit uncommon positioning-related complication in perioperative medicine reported to occur in 0.14% of surgical cases [3]. However, this has been rarely reported with PP in the setting of ARDS. For example, the landmark PROSEVA trial does not even mention PNIs as a complication of PP [4]. Hospitals based in the United States have been incorporating PP into the COVID-19-related ARDS treatment plan at a higher rate than normal. The alarmingly high prevalence of PNI in this cohort of IPR patients (12.9%) not only contributes to post COVID-19 patient burden of disability, but raises the question as to why patients with COVID-19 ARDS are so vulnerable to PNI?

The reason for the high rate of PNI associated with PP in COVID-19 ARDS is likely multifactorial. There were high rates of diabetes mellitus, obesity, and older age seen in our cohort which are characteristics of severe COVID-19-related ARDS patients [5] as well as established risk factors for PNIs [6]. COVID-19 has been associated with muscle injury, acute inflammatory demyelinating polyneuropathy, and a virus-induced state of hyperinflammation and hypercoagulability which may play a role in the vulnerability of peripheral nerves in this patient population [7]. An overlap in COVID-19 mechanisms with type II diabetes mellitus, which itself is associated with neuropathies by a combination of immune, inflammatory, and vascular involvement seems likely [8]. As the subjects described in this study presented from six different tertiary care facilities, hospital specific approaches to PP cannot alone account for these PNIs. Additionally, since only two patients are believed to have received greater than four PP sessions (**Table 1**), it is unlikely that extreme duration of PP was the main cause of these PNIs. To put this in perspective, the average number of PP sessions in PROSEVA trial was 4 [4].

To mitigate the PNI risk associated with PP for COVID-19-related ARDS, careful consideration must be given to protection of the elbow, upper arm and shoulder given the distribution of injuries reported here (**Figure 1b**). Reduction in the mechanical loads on peripheral nerves, specifically avoiding positions of prolonged focal compression and/or stretching of nerves should be an immediate focus. There are undoubtedly lessons to be learned from perioperative medicine [9] to optimize positioning, frequency of repositioning, unloading and cushioning susceptible nerve compression sites, as well as monitoring for early signs of focal injury by visual inspection and physical examination. Ultrasound elastography may prove to be a useful quantitative measurement tool for its sensitivity to increases in nerve tension and stiffness [10]. Limitations of this study include missing clinical data for some patients, the lack of a control group, and its retrospective design which precludes establishment of a causal relationship between PP and PNI in the setting of COVID-19-related ARDS.

In conclusion, PNI after PP for the management of severe COVID-19-related ARDS patients is surprisingly common. Physicians must be aware of this elevated susceptibility to PNI in severe COVID-19 and refined standard PP protocols in order to reduce the risk.

## Data Availability

The de-identified clinical data presented here will be made available to academic-based medical and health researchers on request.

## Abbreviations list

(COVID-19): Coronavirus disease 2019
(ARDS): Acute respiratory distress syndrome
(PP): Prone positioning
(PNI): Peripheral nerve injury
(IPR): Inpatient rehabilitation
(EMG/NCS): Electromyography and nerve conduction studies

